# Effects of human lactoferrin combined with low-dose iron on iron status, hair and skin in women: a randomized, double-blind pilot study

**DOI:** 10.64898/2026.11.25.26368991

**Authors:** Ecaterina Vasenina, Cassandra Evans, Minh Chau, David E Barney, Ross Peterson

**Affiliations:** Human Performance Lab, Department of Health Sciences and Human Performance, The University of Tampa, Tampa, FL, United States; Helaina, Inc., New York, NY, United States; College of Allopathic Medicine, Medicine, Nova Southeastern University, Fort Lauderdale, FL, United States; Pennington Biomedical Research Center, Baton Rouge, LA, United States

**Keywords:** lactoferrin, iron deficiency, ferritin, hair shedding, skin health, hepcidin, pilot randomized trial, women’s health

## Abstract

**Background:** Iron deficiency is common in women of reproductive age, frequently occurs without anemia, and is often accompanied by fatigue and hair shedding. Lactoferrin (LF), an iron-binding glycoprotein, may support iron absorption without the hepcidin response provoked by higher-dose iron, but a human-equivalent LF (heqLF; effera^®^) produced by precision fermentation has not been tested in a randomized trial.

**Objective:** To examine the effects of two doses of heqLF combined with low-dose iron on iron status, inflammation, hair shedding, and participant-reported wellness in women with low ferritin.

**Methods:** In this randomized, double-blind, parallel-group, decentralized pilot study, 31 women aged 18–64 years with ferritin <35 ng/mL were randomized to 100 or 50 mg/d of heqLF, each with 5 mg/d of iron as ferrous bisglycinate, for 6 weeks; 27 were included in the modified intention-to-treat analysis. Iron, hematological, and inflammatory biomarkers were measured at Weeks 0, 2, 4, and 6, and hair shedding (self-administered modified hair pull test) and participant-reported outcomes were assessed weekly.

**Results:** Ferritin, the primary endpoint, did not change in either group (100 mg: +0.9%; 50 mg: +11.3%; both P ≥ 0.34). With both groups combined, total iron-binding capacity decreased by 17 µg/dL, unsaturated iron-binding capacity by 34 µg/dL, and transferrin by 13 mg/dL (all adjusted P ≤ 0.028), whereas increases in serum iron and transferrin saturation were not statistically significant in the primary analysis. High-sensitivity C-reactive protein and hemoglobin did not change. Hair pull counts decreased by 62% with 100 mg (adjusted P = 0.017) and by 39% with 50 mg (adjusted P = 0.039). Participant-reported wellness and hair scores improved in both groups, and skin scores improved with 100 mg only. No outcome differed between doses.

**Conclusions:** In women with low ferritin, 6 weeks of heqLF with low-dose iron was accompanied by lower iron-binding capacity and transferrin, consistent with improved iron availability, as well as reduced hair shedding and improved wellness, without a change in ferritin. Because the study had no placebo or iron-only control arm, these exploratory findings warrant confirmation in a larger placebo-controlled trial.

**Clinical trial registration:** www.clinicaltrials.gov, identifier NCT07953972

## 1 Introduction

Iron deficiency (ID) is arguably the most prevalent micronutrient deficiency worldwide and disproportionately affects women of reproductive age as a consequence of menstrual blood loss, pregnancy-related demands, and frequently inadequate dietary iron intake (1, 2). ID develops along a continuum: depletion of storage iron, reflected by a low serum ferritin concentration, precedes a decline in circulating iron (lower serum iron and transferrin saturation), iron-deficient erythropoiesis and, ultimately, iron-deficiency anemia (IDA) (1). Even in the absence of anemia, ID can impair work capacity, cognitive function, and quality of life, and it is commonly accompanied by fatigue and changes in hair, skin, and nail quality (3–6). Because anemia is a late manifestation, a substantial proportion of women with depleted iron stores remain unrecognized when screening relies on hemoglobin alone (3). In a nationally representative analysis of US National Health and Nutrition Examination Survey (NHANES) data, 38.6% of females aged 12–21 years had ID (ferritin <25 µg/L), and in 83.6% of affected individuals ID occurred without anemia (7).

Non-anemic ID is increasingly recognized as a clinically meaningful condition rather than a benign laboratory finding (3, 4). In randomized controlled trials, iron supplementation reduced fatigue in non-anemic women with low ferritin (8, 9), and a systematic review of randomized trials in non-anemic iron-deficient adults found that iron supplementation reduced subjective fatigue, although objective measures of physical capacity did not improve (10). Restoration of iron status has also been associated with improved attention, memory, and learning in young women (5). These observations are particularly relevant to physically active women. Iron is central to oxygen transport and oxidative metabolism, and ID is commonly diagnosed in athletes, particularly females and endurance athletes (11). Exercise-related mechanisms, including hemolysis, gastrointestinal and sweat iron losses, and transient post-exercise elevations in hepcidin driven by interleukin-6 (IL-6), may compound menstrual losses and low energy availability (12, 13). In iron-deficient non-anemic endurance athletes, iron treatment improves iron status and aerobic capacity (14).

Hair shedding is among the most frequently reported, yet least systematically studied, consequences of low iron status. Hair follicle matrix cells are among the most rapidly dividing cells in the body and depend on iron-containing enzymes, including ribonucleotide reductase, the rate-limiting enzyme of DNA synthesis, providing a biological rationale for an association between iron depletion and telogen hair shedding (6). Several case-control studies have reported lower ferritin concentrations in women with diffuse non-scarring hair loss (15–18). The evidence is not uniform, however; in a controlled study of 381 women with female pattern hair loss or chronic telogen effluvium, ID was common but not more prevalent than in controls (19), and others have found no association between ferritin concentrations above 10 µg/L and hair loss activity (20). Interventional data remain limited (6, 21). Iron-independent pathways may also be relevant: lactoferrin (LF), an iron-binding glycoprotein described below, promoted hair growth in mice and increased the proliferation of dermal papilla cells, which regulate the hair growth cycle, through ERK/Akt and Wnt signaling pathways (22), although these findings derive from animal and cell culture models using topical or parenteral administration at supraphysiological concentrations. The potential roles of LF in hair and skin health have recently been reviewed (23). Clinically, active shedding is commonly evaluated using the hair pull test, a simple, standardized assessment (24); adaptation of this test for self-administration may permit repeated monitoring of hair shedding in remote or decentralized trials.

Oral iron salts remain first-line therapy for ID but have important limitations. Systemic iron balance is governed by the hepatic peptide hormone hepcidin, which circulates to bind the cellular iron exporter ferroportin on target tissues, which occludes ferroportin and induces its internalization and degradation, thereby restricting export of absorbed dietary iron from enterocytes, recycled iron from macrophages, and tissue stores in the liver (25–27). Hepcidin primarily responds to iron loading as a negative feedback. In iron-depleted women, a single oral iron dose of ≥60 mg increases serum hepcidin for approximately 24 h, reducing fractional absorption from doses given later the same or next day; consequently, alternate-day dosing maximizes fractional absorption (28, 29). Inflammation also signals for hepcidin. As such, sources of inflammation including infection, strenuous exercise, and excess adiposity also elevate hepcidin and limits the effectiveness of supplemental iron (26, 30). Oral iron salts also frequently cause gastrointestinal side effects, which undermine adherence (31), and unabsorbed iron reaching the colon may shift the gut microbiota toward potentially pathogenic taxa and promote intestinal inflammation (32, 33). Collectively, these constraints have motivated interest in adjuncts that improve iron absorption rather than simply increasing the amount of iron delivered.

LF is an ∼80 kDa iron-binding glycoprotein of the transferrin family that reversibly binds two ferric ions with high affinity and is abundant in human milk, exocrine secretions such as saliva, tears, and bile, and neutrophil secondary granules (34, 35). Several mechanisms may allow LF to support iron homeostasis. First, LF is recognized by a specific intestinal LF receptor (intelectin-1) and internalized via clathrin-mediated endocytosis independently of its iron saturation, providing a route for receptor-mediated uptake (36, 37). Second, LF exerts anti-inflammatory activity relevant to iron regulation: in human monocytes/macrophages, LF prevented the lipopolysaccharide-induced decrease in ferroportin by reducing IL-6 secretion (38), and clinical studies have linked LF-associated improvements in iron status to reductions in circulating IL-6 (39), although these effects have been reported mainly in specific subpopulations, and a large recent trial found no meaningful change in inflammatory markers with bLF (40). These effects may mitigate inflammatory inhibition of dietary iron aborption and tissue iron export to enhance fractional iron absorption and support iron availability from storage. . Third, LF possesses broader immunomodulatory properties that may be relevant in individuals with low-grade inflammation (41).

Clinical studies, predominantly using bovine LF (bLF), support these mechanisms. In pregnant women with ID and IDA, oral bLF increased red blood cell count, hemoglobin, serum iron, and ferritin while decreasing serum IL-6, whereas ferrous sulfate increased IL-6 and failed to improve hematological parameters (42); similar benefits were observed in non-pregnant women of childbearing age receiving bLF 100 mg twice daily (42). A meta-analysis of four randomized trials in 600 pregnant women found that bLF was as effective as ferrous sulfate in improving hematological parameters, with significantly fewer gastrointestinal side effects (43), and a broader meta-analysis reported greater improvements in serum iron, ferritin, and hemoglobin and lower IL-6 with LF than with ferrous sulfate (44). Notably, in a study of 16 female long-distance runners, 8 weeks of LF plus iron maintained ferritin, serum iron, and red blood cell count during summer training, whereas these indices declined with the same iron dose (5 mg ferric pyrophosphate) alone (45), suggesting that LF may complement conventional iron supplementation in active women. Not all trials have been positive, however. In a randomized trial of 555 Bangladeshi women with IDA, bLF at 200 or 400 mg/d for 12 weeks, given without iron, was inferior to ferrous sulfate (60 mg/d elemental iron) for improving hemoglobin and produced only small increases in ferritin, although constipation and black stools were less frequent (40); the authors suggested that LF may be more useful as an adjunct to iron than as a substitute.

Human and bovine LF share only ∼69% amino acid sequence identity and differ in glycosylation (46). Although bLF can be taken up by the human intestinal LF receptor (35), human LF is its native ligand (47). Until recently, however, human LF could not be produced at a scale suitable for supplementation. Precision fermentation has enabled the production of a human-equivalent LF (heqLF; effera^®^, Helaina Inc.) produced by the yeast *Komagataella phaffii*, which has an amino acid sequence identical to that of native human milk LF and equivalent secondary and tertiary structures (48). The safety of heqLF has been evaluated following an expert panel-guided roadmap (49), which included an allergenicity assessment (50), a 4-week immunotoxicity study in rats (51), an in vitro digestion study under physiologically relevant conditions (52), and a randomized, double-blind, controlled trial in which 66 healthy adults received heqLF at 0.34 or 3.4 g/d or bLF at 3.4 g/d for 28 days to evaluate alloimmunization potential (53). On the basis of these data, heqLF meets the generally recognized as safe (GRAS) standard. Emerging clinical data also suggest benefits for iron status: in a recent case series of two active women with persistently low iron and poor response to, or intolerance of, standard oral iron, 100 mg/d of heqLF for 6–20 weeks was followed by notable increases in ferritin, with one participant also showing marked increases in serum iron and transferrin saturation, alongside improved energy and exercise capacity and no gastrointestinal complaints (54). However, no randomized controlled trial has examined heqLF administered with iron in women with low ferritin, nor its effects on commonly reported outcomes such as fatigue, mood, and hair shedding.

Therefore, the purpose of this randomized, double-blind, parallel-group pilot study was to examine the effects of two doses of heqLF (50 mg/d or 100 mg/d, each with 5 mg/d iron), administered for 6 weeks, on (1) biomarkers of iron status (ferritin, serum iron, transferrin, total and unsaturated iron-binding capacity, and transferrin saturation) and hematological indices; (2) systemic inflammation (high-sensitivity C-reactive protein); (3) hair shedding, self-assessed using a modified hair pull test; and (4) participant-reported wellness (energy, fatigue, and mood) and perceived hair and skin health in adult women with low ferritin concentrations. We hypothesized that both doses would improve biomarkers of iron status, reduce hair shedding, and improve participant-reported wellness over 6 weeks, and that these effects would be greater with the higher LF dose.

## 2 Materials and methods

### 2.1 Study design

This pilot, proof-of-concept study used a randomized, double-blind, parallel-group design to examine the effects of two doses of heqLF administered with iron on indices of iron status, inflammation, hair shedding, and subjective measures of wellness in adult women with low ferritin concentrations. Eligible participants were women aged 18–64 years with a documented ferritin concentration <35 ng/mL within the previous 6 months and a body mass index (BMI) between 18.5 and 32.5 kg/m². The ferritin criterion used in the present study (<35 ng/mL) lies slightly above the physiologically based threshold for iron-deficient erythropoiesis (55) and therefore captures women ranging from those at risk of functional iron deficiency to those with absolute iron deficiency by the WHO criterion (56).

Individuals were excluded if they had gastrointestinal disorders that could affect study outcomes, select chronic medical conditions, acute illness or infection within the previous 30 days, or recent changes in medications that could confound study outcomes, or if they were pregnant or lactating. Participants were supplemented for 6 weeks, with blood sampling, hair pull tests, and questionnaires at baseline (Week 0) and at regular intervals through Week 6 (Section 2.3; Figure 1A). No placebo or iron-only control arm was included.

**Figure 1.**
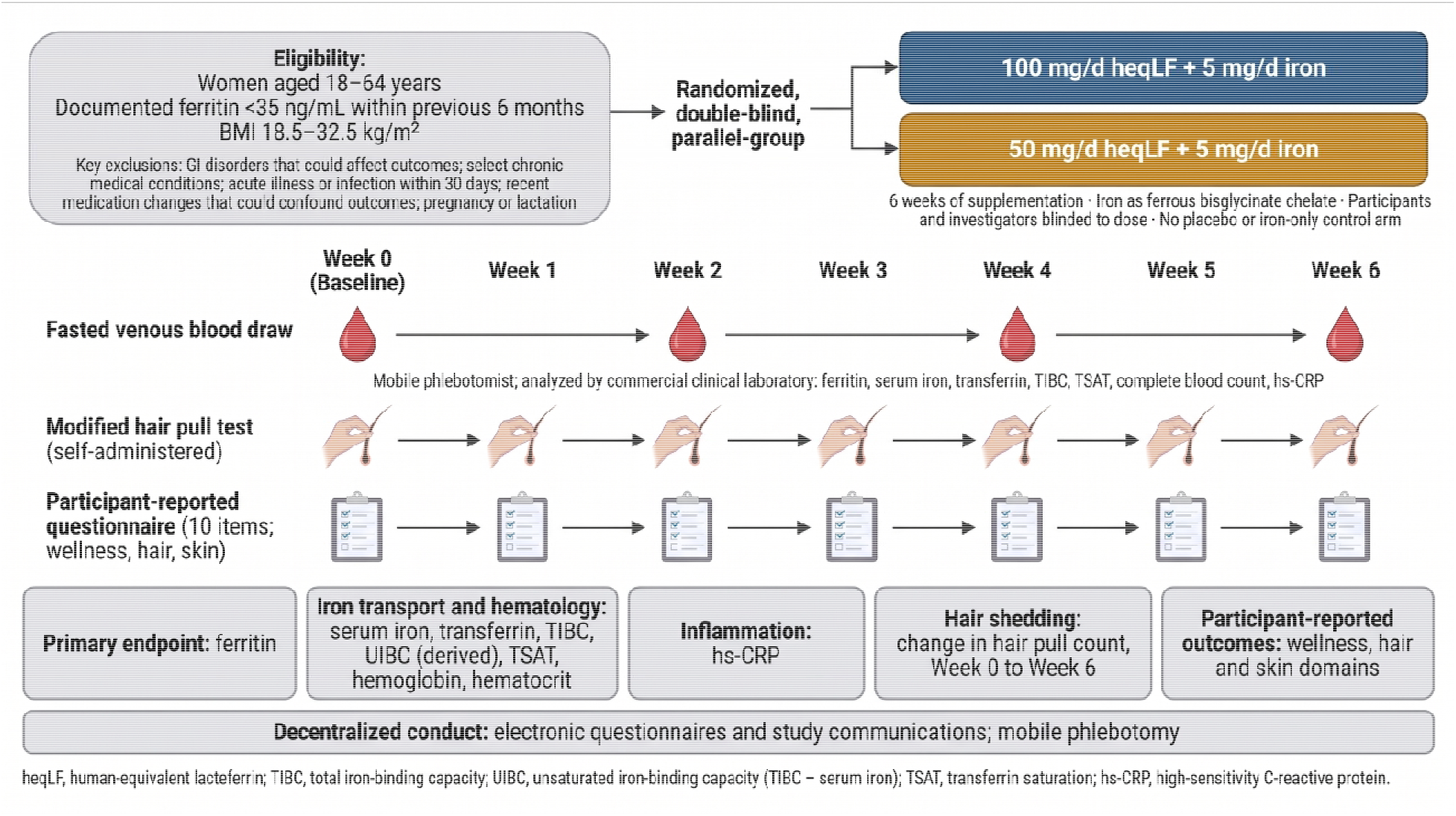
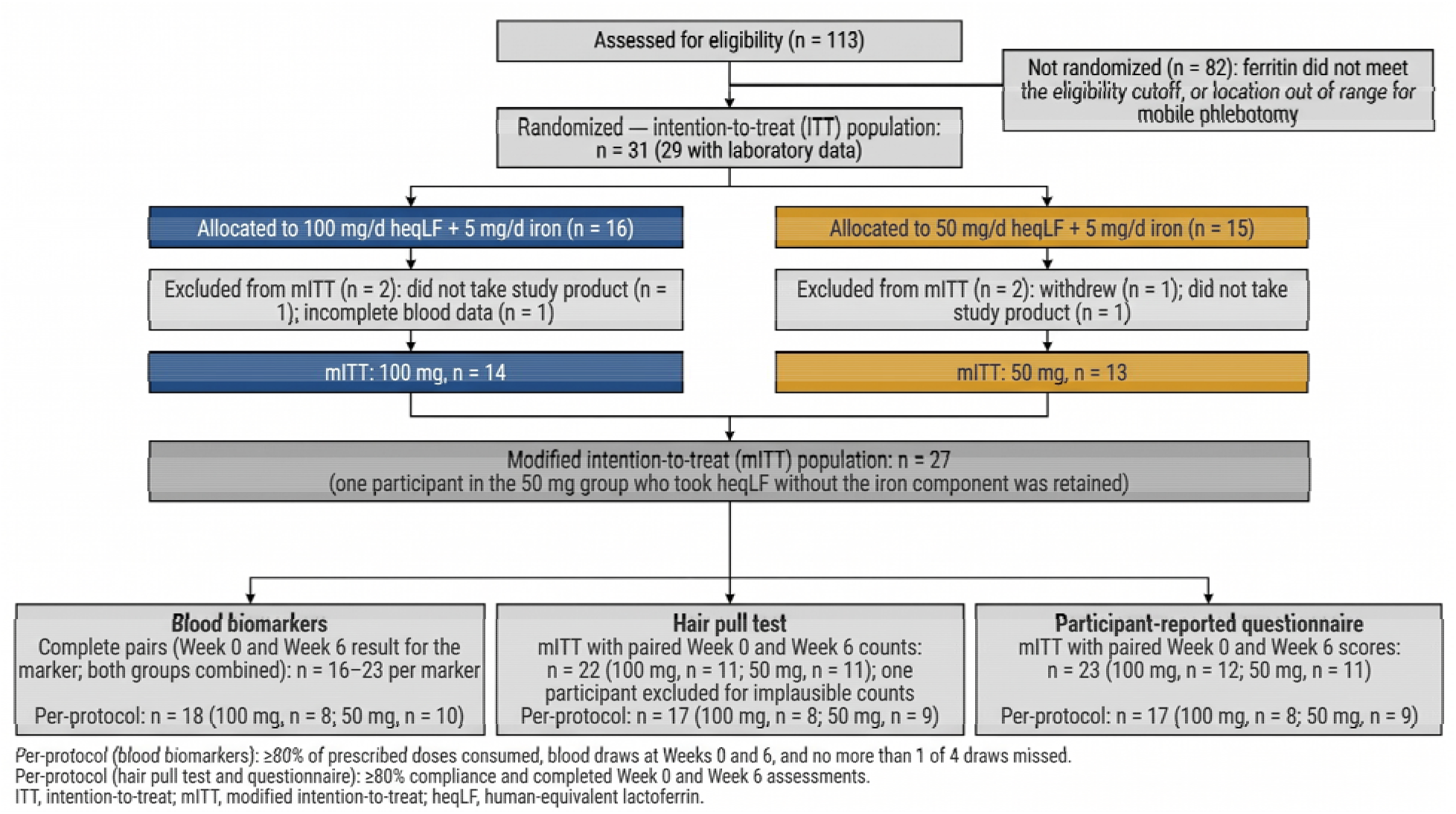
Study design and participant flow. (A) Study design, interventions, and assessment schedule. (B) Participant flow and analysis populations.

All participants provided informed consent in accordance with a protocol approved by the BRANY Institutional Review Board (Protocol Version 2; approved May 8, 2026). The study was registered at ClinicalTrials.gov ([NCT07953972]).

### 2.2 Supplementation protocol

Participants were randomly assigned to receive either 100 mg/d of heqLF (effera^®^, Helaina Inc.) plus 5 mg/d of iron or 50 mg/d of heqLF plus 5 mg/d of iron for 6 weeks. Iron was provided as ferrous bisglycinate chelate (Ferrochel®, Albion Laboratories; Iron 5 mg Bio-Max Series, Kirkman Group, Lake Oswego, OR, USA). Both participants and investigators were blinded to dose allocation. Compliance was assessed by pill count and expressed as the percentage of prescribed doses consumed; participants consuming ≥80% of prescribed doses were considered compliant, and participants without a reported pill count were not excluded as non-compliant.

### 2.3 Study procedures

The study was conducted in a decentralized manner: questionnaires and study communications were completed electronically, and fasted venous blood samples were collected by trained mobile phlebotomy professionals. Participants completed baseline demographic and health assessments, including weekly hours of resistance training and cardiovascular exercise, self-reported duration of low iron, and current symptoms. The screening ferritin value used to determine eligibility was the documented result obtained within the 6 months before enrollment. The modified hair pull test and the participant-reported outcome questionnaire were completed at baseline (Week 0) and weekly through Week 6.

Venous blood samples were collected at Weeks 0, 2, 4, and 6 and analyzed by a commercial clinical laboratory (Labcorp) for ferritin, serum iron, transferrin, total iron-binding capacity (TIBC), transferrin saturation (TSAT), complete blood count, and high-sensitivity C-reactive protein (hs-CRP). Iron indices and hs-CRP were measured in serum, and the complete blood count in EDTA whole blood. Serum iron and TSAT were measured in two panels: an iron and TIBC panel (Fe panel), in which TSAT was calculated as (serum iron ÷ TIBC) × 100; and a transferrin panel (Tf panel), in which TSAT was calculated as (serum iron ÷ Tf) × 70.9. Unsaturated iron-binding capacity (UIBC) was derived from the Fe panel as TIBC − serum iron and is therefore not independent of these two measures. Fe panel values are reported in Table 3; Tf panel values are reported in Table 4.

**Table 1.** Baseline characteristics of the modified intention-to-treat population.

| **Variable** | **Total (n = 27)** | **100 mg (n = 14)** | **50 mg (n = 13)** | **P** |
|---|---|---|---|---|
| *Participant characteristics* |  |  |  |  |
| Age (years) | 36.9 ± 9.0 | 34.4 ± 7.4 | 39.8 ± 10.1 | 0.14 |
| Height (cm) | 164.5 ± 7.9 | 165.3 ± 9.2 | 163.6 ± 6.4 | 0.60 |
| Weight (kg) | 69.3 ± 13.4 | 67.3 ± 11.5 | 71.7 ± 15.5 | 0.43 |
| BMI (kg/m<sup>2</sup>) | 25.6 ± 4.6 | 24.6 ± 3.8 | 26.7 ± 5.4 | 0.28 |
| Resistance training (h/wk) | 2.2 ± 2.2 | 2.3 ± 2.1 | 2.2 ± 2.4 | 0.90 |
| Cardiovascular exercise (h/wk) | 4.6 ± 3.3 | 4.6 ± 2.2 | 4.7 ± 4.3 | 0.95 |
| Duration of low iron status, n (%) |  |  |  | 0.26 |
| Less than 3 months | 1 (3.8) | 1 (7.1) | 0 (0.0) |  |
| 3–6 months | 2 (7.7) | 2 (14.3) | 0 (0.0) |  |
| 6–12 months | 4 (15.4) | 3 (21.4) | 1 (8.3) |  |
| More than 1 year | 19 (73.1) | 8 (57.1) | 11 (91.7) |  |
| *Iron storage* |  |  |  |  |
| Screening ferritin (ng/mL) | 14.0 ± 9.0 | 14.3 ± 10.0 | 13.6 ± 8.1 | 0.83 |
| <30 ng/mL, n (%) | 25 (92.6) | 12 (85.7) | 13 (100.0) | 0.48 |
| <25 ng/mL, n (%) | 24 (88.9) | 12 (85.7) | 12 (92.3) | 1.00 |
| <15 ng/mL, n (%) | 17 (63.0) | 9 (64.3) | 8 (61.5) | 1.00 |
| Week 0 ferritin (ng/mL) | 31.8 ± 17.6 | 30.4 ± 17.4 | 33.4 ± 18.5 | 0.68 |
| <30 ng/mL, n (%) | 12 (46.2) | 8 (57.1) | 4 (33.3) | 0.27 |
| <25 ng/mL, n (%) | 9 (34.6) | 5 (35.7) | 4 (33.3) | 1.00 |
| <15 ng/mL, n (%) | 7 (26.9) | 3 (21.4) | 4 (33.3) | 0.67 |
| *Iron transport (Week 0)* |  |  |  |  |
| Serum iron, Fe panel (µg/dL) | 71.7 ± 38.4 | 78.7 ± 42.9 | 63.4 ± 32.2 | 0.31 |
| Serum iron, Tf panel (µg/dL) | 80.0 ± 40.3 | 86.8 ± 42.0 | 69.8 ± 38.0 | 0.36 |
| TSAT, Fe panel (%) | 21.3 ± 12.8 | 23.1 ± 14.1 | 19.2 ± 11.4 | 0.45 |
| TSAT, Tf panel (%) | 20.5 ± 11.4 | 21.8 ± 11.8 | 18.6 ± 11.2 | 0.56 |
| Transferrin (mg/dL) | 291.1 ± 43.2 | 294.8 ± 49.3 | 285.8 ± 34.5 | 0.62 |
| TIBC (µg/dL) | 357.2 ± 53.1 | 361.1 ± 55.3 | 352.7 ± 52.4 | 0.69 |
| UIBC (µg/dL) | 285.6 ± 79.2 | 282.4 ± 83.7 | 289.2 ± 77.0 | 0.83 |
| *Functional iron (Week 0)* |  |  |  |  |
| Hemoglobin (g/dL) | 13.0 ± 1.1 | 12.8 ± 1.5 | 13.1 ± 0.7 | 0.62 |
| Anemia, n (%)† | 3 (12.5) | 2 (16.7) | 1 (8.3) | 1.00 |
| Iron-deficiency anemia, n (%)‡ | 3 (12.5) | 2 (16.7) | 1 (8.3) | 1.00 |
| Hematocrit (%) | 40.2 ± 2.8 | 39.9 ± 3.4 | 40.5 ± 2.1 | 0.58 |
| MCV (fL) | 89.5 ± 6.1 | 88.1 ± 6.2 | 91.0 ± 5.9 | 0.25 |
| MCH (pg) | 28.9 ± 2.6 | 28.3 ± 3.0 | 29.4 ± 2.1 | 0.33 |
| RDW (%) | 13.7 ± 2.1 | 13.3 ± 1.7 | 14.1 ± 2.5 | 0.40 |
| *Inflammation (Week 0)* |  |  |  |  |
| hs-CRP (mg/L) | 2.11 ± 2.25 | 2.46 ± 2.90 | 1.73 ± 1.25 | 0.44 |
| WBC (×10<sup>3</sup>/µL) | 5.9 ± 2.1 | 6.0 ± 2.1 | 5.8 ± 2.1 | 0.81 |
| Platelets (×10<sup>3</sup>/µL) | 301 ± 82 | 314 ± 84 | 288 ± 81 | 0.44 |

Values are mean ± SD or n (%), with percentages calculated from participants with non-missing data. Groups were compared with Welch's unpaired two-sided *t*-tests (continuous variables) or Fisher's exact tests (categorical variables). Laboratory values are from Week 0 except screening ferritin, which was obtained before enrollment. †Hemoglobin <12.0 g/dL. ‡Hemoglobin <12.0 g/dL with ferritin <15 ng/mL (WHO criteria). Duration of low iron status was self-reported and not defined by a ferritin cutoff. Participants with missing data, total (100 mg/50 mg): height, weight, BMI, training habits, and duration of low iron status, 1 (0/1); Week 0 ferritin, Fe panel serum iron and TSAT, TIBC, and UIBC, 1 (0/1); Tf panel serum iron and TSAT, 7 (2/5); transferrin, 5 (1/4); hemoglobin, hematocrit, MCV, MCH, RDW, WBC, and platelets, 3 (2/1); hs-CRP, 4 (2/2). Abbreviations: BMI, body mass index; Fe panel, iron and TIBC panel; hs-CRP, high-sensitivity C-reactive protein; MCH, mean corpuscular hemoglobin; MCV, mean corpuscular volume; RDW, red cell distribution width; Tf panel, transferrin panel; TIBC, total iron-binding capacity; TSAT, transferrin saturation; UIBC, unsaturated iron-binding capacity; WBC, white blood cell count.

**Table 2.** Frequency of reported symptoms at baseline.

| **Symptom** | **n** | **%** |
|---|---|---|
| Fatigue | 25 | 100.0 |
| Brain fog | 22 | 88.0 |
| Hair shedding/thinning | 21 | 84.0 |
| Brittle nails | 13 | 52.0 |
| Dizziness | 11 | 44.0 |
| Restless legs | 10 | 40.0 |
| Shortness of breath | 8 | 32.0 |
| Other | 2 | 8.0 |

n = 25 participants with symptom data. Participants could endorse more than one symptom; therefore, percentages do not sum to 100.

**Table 3.** Blood biomarkers at Week 0 and Week 6 and change by dose group (modified intention-to-treat).

| Marker / group | n (Wk 0/6) | Week 0 | Week 6 | Δ Week 0→6 (95% CI) | P | Adj. P (×3) |
|---|---|---|---|---|---|---|
| **Ferritin (ng/mL)†** |  |  |  |  |  |  |
| 100 mg | 14/11 | 25.4 (17.9, 35.8) | 25.6 (17.9, 36.5) | +0.9% (−18.4, +24.8) | 0.93 | 1.00 |
| 50 mg | 12/10 | 27.5 (18.9, 40.0) | 30.6 (20.9, 44.9) | +11.3% (−11.1, +39.4) | 0.34 | 1.00 |
| 50 − 100 mg |  | — | — | +10.3% (−19.0, +50.3) | 0.53 | 1.00 |
| Pooled | 26 | — | — | +5.7% (−9.1, +22.9) | 0.46 | 1.00 |
| **BRINDA-adjusted ferritin (ng/mL)†** |  |  |  |  |  |  |
| 100 mg | 14/11 | 19.5 (13.7, 27.9) | 19.3 (13.4, 27.8) | −1.2% (−20.7, +23.1) | 0.91 | 1.00 |
| 50 mg | 12/10 | 21.4 (14.6, 31.4) | 24.2 (16.3, 35.8) | +12.9% (−10.5, +42.4) | 0.30 | 0.90 |
| 50 − 100 mg |  | — | — | +14.2% (−17.0, +57.2) | 0.41 | 1.00 |
| Pooled | 26 | — | — | +5.2% (−10.0, +23.0) | 0.52 | 1.00 |
| **Serum iron (µg/dL)** |  |  |  |  |  |  |
| 100 mg | 14/12 | 79 (58, 100) | 91 (69, 114) | +13 (−11, +37) | 0.29 | 0.88 |
| 50 mg | 12/11 | 63 (40, 85) | 78 (55, 101) | +15 (−10, +41) | 0.24 | 0.71 |
| 50 − 100 mg |  | — | — | +2 (−33, +38) | 0.89 | 1.00 |
| Pooled | 27 | — | — | +14 (−4, +32) | 0.12 | 0.37 |
| **TSAT (%)** |  |  |  |  |  |  |
| 100 mg | 14/12 | 23.1 (16.1, 30.0) | 27.7 (20.4, 35.0) | +4.6 (−2.3, +11.6) | 0.19 | 0.56 |
| 50 mg | 12/11 | 18.9 (11.5, 26.3) | 24.2 (16.7, 31.7) | +5.2 (−2.2, +12.6) | 0.16 | 0.48 |
| 50 − 100 mg |  | — | — | +0.6 (−9.5, +10.7) | 0.91 | 1.00 |
| Pooled | 27 | — | — | +5.0 (−0.2, +10.1) | 0.060 | 0.18 |
| **Transferrin (mg/dL)** |  |  |  |  |  |  |
| 100 mg | 13/12 | 299 (275, 323) | 284 (259, 308) | −15 (−28, −2) | **0.020** | 0.061 |
| 50 mg | 9/10 | 281 (256, 307) | 272 (247, 298) | −9 (−24, +7) | 0.26 | 0.78 |
| 50 − 100 mg |  | — | — | +6 (−14, +26) | 0.54 | 1.00 |
| Pooled | 27 | — | — | −13 (−22, −3) | **0.009** | **0.028** |
| **TIBC (µg/dL)** |  |  |  |  |  |  |
| 100 mg | 14/12 | 361 (332, 390) | 342 (314, 370) | −19 (−32, −6) | **0.005** | **0.014** |
| 50 mg | 12/11 | 350 (319, 380) | 335 (306, 364) | −15 (−29, 0) | **0.045** | 0.14 |
| 50 − 100 mg |  | — | — | +5 (−15, +24) | 0.62 | 1.00 |
| Pooled | 27 | — | — | −17 (−27, −8) | **<0.001** | **0.002** |
| **UIBC (µg/dL)‡** |  |  |  |  |  |  |
| 100 mg | 14/12 | 282 (241, 324) | 249 (207, 292) | −33 (−61, −6) | **0.019** | 0.058 |
| 50 mg | 12/11 | 289 (245, 333) | 254 (210, 298) | −35 (−65, −5) | **0.022** | 0.065 |
| 50 − 100 mg |  | — | — | −2 (−42, +39) | 0.93 | 1.00 |
| Pooled | 27 | — | — | −34 (−55, −14) | **0.001** | **0.004** |
| **Hemoglobin (g/dL)** |  |  |  |  |  |  |
| 100 mg | 12/12 | 12.8 (12.1, 13.5) | 12.9 (12.2, 13.5) | +0.1 (−0.3, +0.4) | 0.79 | 1.00 |
| 50 mg | 12/12 | 12.9 (12.2, 13.6) | 13.0 (12.3, 13.6) | +0.1 (−0.3, +0.4) | 0.76 | 1.00 |
| 50 − 100 mg |  | — | — | +0.0 (−0.5, +0.5) | 0.98 | 1.00 |
| Pooled | 26 | — | — | +0.1 (−0.2, +0.3) | 0.65 | 1.00 |
| **Hematocrit (%)** |  |  |  |  |  |  |
| 100 mg | 12/12 | 39.9 (38.2, 41.5) | 40.3 (38.7, 41.9) | +0.4 (−0.9, +1.7) | 0.51 | 1.00 |
| 50 mg | 12/12 | 40.1 (38.5, 41.8) | 40.3 (38.7, 41.9) | +0.1 (−1.1, +1.4) | 0.81 | 1.00 |
| 50 − 100 mg |  | — | — | −0.3 (−2.1, +1.5) | 0.77 | 1.00 |
| Pooled | 26 | — | — | +0.3 (−0.6, +1.2) | 0.51 | 1.00 |
| **hs-CRP (mg/L)†** |  |  |  |  |  |  |
| 100 mg | 12/11 | 1.25 (0.68, 2.29) | 1.18 (0.63, 2.21) | −5.2% (−44.1, +60.9) | 0.84 | 1.00 |
| 50 mg | 11/11 | 1.21 (0.64, 2.28) | 0.99 (0.53, 1.87) | −17.7% (−52.4, +42.3) | 0.48 | 1.00 |
| 50 − 100 mg |  | — | — | −13.2% (−59.5, +85.8) | 0.71 | 1.00 |
| Pooled | 27 | — | — | −11.1% (−39.2, +30.0) | 0.54 | 1.00 |
| **WBC (×10³/µL)†** |  |  |  |  |  |  |
| 100 mg | 12/12 | 5.78 (4.78, 6.98) | 5.92 (5.19, 6.73) | +2.4% (−11.4, +18.3) | 0.74 | 1.00 |
| 50 mg | 12/12 | 5.62 (4.65, 6.79) | 5.37 (4.72, 6.12) | −4.4% (−17.2, +10.4) | 0.53 | 1.00 |
| 50 − 100 mg |  | — | — | −6.6% (−23.9, +14.5) | 0.50 | 1.00 |
| Pooled | 26 | — | — | −0.8% (−10.3, +9.6) | 0.86 | 1.00 |

*Note.* Values are estimated marginal means (95% CI) from linear mixed models (group, visit, and group × visit; REML; Kenward–Roger degrees of freedom). Rows labeled 50 − 100 mg give the between-group difference in change. Pooled changes are from models without a group term (n = participants contributing). P, unadjusted P value; Adj. P (×3), P value after Bonferroni adjustment across the three post-baseline time points. P-values <0.05 are shown in bold. Serum iron and TSAT are from the Fe panel. †Log-transformed; values are geometric means and Δ is percentage change. ‡UIBC = TIBC − serum iron (derived). Abbreviations: BRINDA, Biomarkers Reflecting Inflammation and Nutritional Determinants of Anemia; hs-CRP, high-sensitivity C-reactive protein; TIBC, total iron-binding capacity; TSAT, transferrin saturation; UIBC, unsaturated iron-binding capacity; WBC, white blood cell count.

**Table 4.**
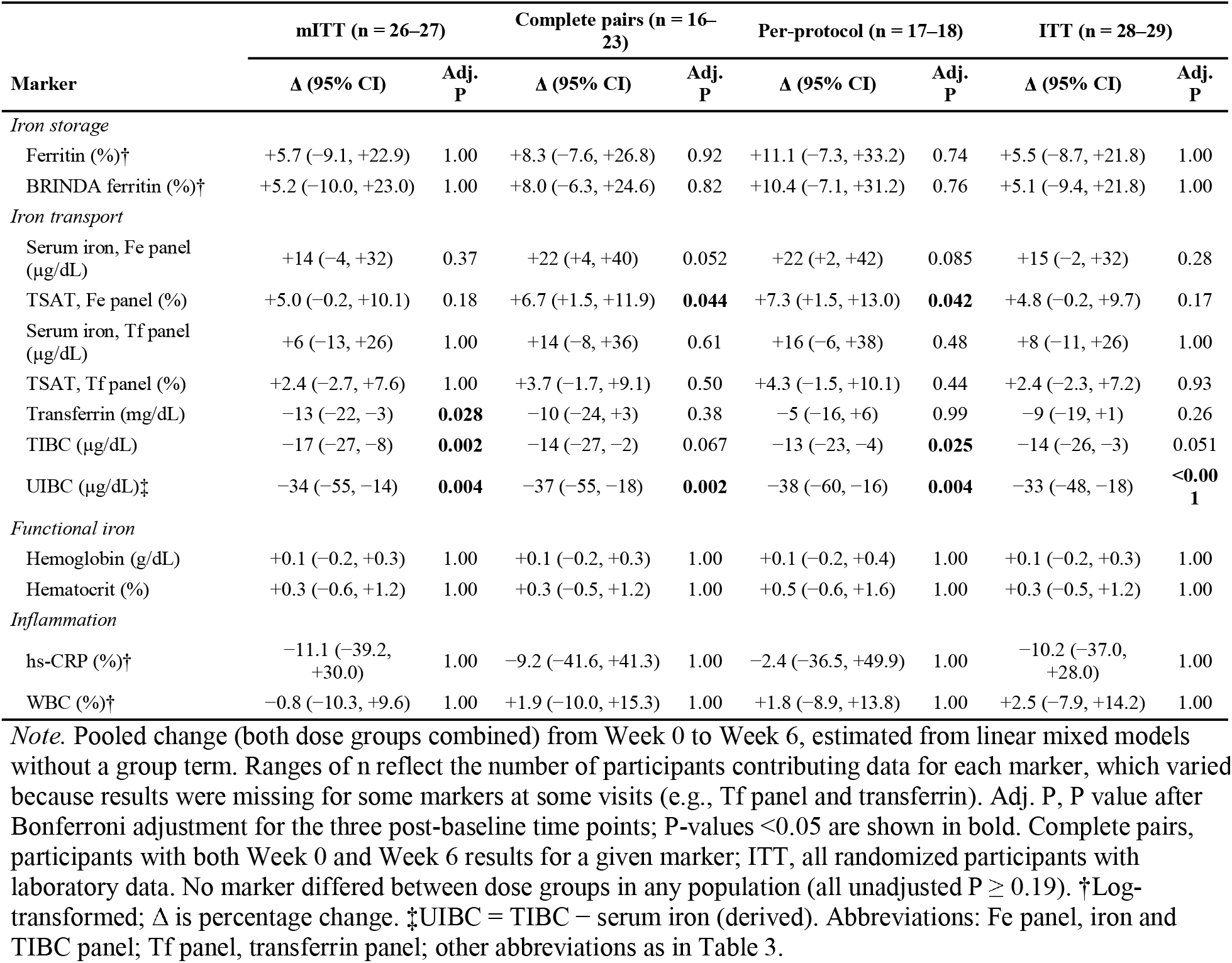
Pooled change in blood biomarkers from Week 0 to Week 6 across analysis populations.

|  | **mITT (n = 26–27)** |  | **Complete pairs (n = 16–23)** |  | **Per-protocol (n = 17–18)** |  | **ITT (n = 28–29)** |  |
|---|---|---|---|---|---|---|---|---|
| **Marker** | **Δ (95% CI)** | **Adj. P** | **Δ (95% CI)** | **Adj. P** | **Δ (95% CI)** | **Adj. P** | **Δ (95% CI)** | **Adj. P** |
| *Iron storage* |  |  |  |  |  |  |  |  |
| Ferritin (%)† | +5.7 (−9.1, +22.9) | 1.00 | +8.3 (−7.6, +26.8) | 0.92 | +11.1 (−7.3, +33.2) | 0.74 | +5.5 (−8.7, +21.8) | 1.00 |
| BRINDA ferritin (%)† | +5.2 (−10.0, +23.0) | 1.00 | +8.0 (−6.3, +24.6) | 0.82 | +10.4 (−7.1, +31.2) | 0.76 | +5.1 (−9.4, +21.8) | 1.00 |
| *Iron transport* |  |  |  |  |  |  |  |  |
| Serum iron, Fe panel (µg/dL) | +14 (−4, +32) | 0.37 | +22 (+4, +40) | 0.052 | +22 (+2, +42) | 0.085 | +15 (−2, +32) | 0.28 |
| TSAT, Fe panel (%) | +5.0 (−0.2, +10.1) | 0.18 | +6.7 (+1.5, +11.9) | **0.044** | +7.3 (+1.5, +13.0) | **0.042** | +4.8 (−0.2, +9.7) | 0.17 |
| Serum iron, Tf panel (µg/dL) | +6 (−13, +26) | 1.00 | +14 (−8, +36) | 0.61 | +16 (−6, +38) | 0.48 | +8 (−11, +26) | 1.00 |
| TSAT, Tf panel (%) | +2.4 (−2.7, +7.6) | 1.00 | +3.7 (−1.7, +9.1) | 0.50 | +4.3 (−1.5, +10.1) | 0.44 | +2.4 (−2.3, +7.2) | 0.93 |
| Transferrin (mg/dL) | −13 (−22, −3) | **0.028** | −10 (−24, +3) | 0.38 | −5 (−16, +6) | 0.99 | −9 (−19, +1) | 0.26 |
| TIBC (µg/dL) | −17 (−27, −8) | **0.002** | −14 (−27, −2) | 0.067 | −13 (−23, −4) | **0.025** | −14 (−26, −3) | 0.051 |
| UIBC (µg/dL)‡ | −34 (−55, −14) | **0.004** | −37 (−55, −18) | **0.002** | −38 (−60, −16) | **0.004** | −33 (−48, −18) | **<0.001** |
| *Functional iron* |  |  |  |  |  |  |  |  |
| Hemoglobin (g/dL) | +0.1 (−0.2, +0.3) | 1.00 | +0.1 (−0.2, +0.3) | 1.00 | +0.1 (−0.2, +0.4) | 1.00 | +0.1 (−0.2, +0.3) | 1.00 |
| Hematocrit (%) | +0.3 (−0.6, +1.2) | 1.00 | +0.3 (−0.5, +1.2) | 1.00 | +0.5 (−0.6, +1.6) | 1.00 | +0.3 (−0.5, +1.2) | 1.00 |
| *Inflammation* |  |  |  |  |  |  |  |  |
| hs-CRP (%)† | −11.1 (−39.2, +30.0) | 1.00 | −9.2 (−41.6, +41.3) | 1.00 | −2.4 (−36.5, +49.9) | 1.00 | −10.2 (−37.0, +28.0) | 1.00 |
| WBC (%)† | −0.8 (−10.3, +9.6) | 1.00 | +1.9 (−10.0, +15.3) | 1.00 | +1.8 (−8.9, +13.8) | 1.00 | +2.5 (−7.9, +14.2) | 1.00 |

*Note.* Pooled change (both dose groups combined) from Week 0 to Week 6, estimated from linear mixed models without a group term. Ranges of n reflect the number of participants contributing data for each marker, which varied because results were missing for some markers at some visits (e.g., Tf panel and transferrin). Adj. P, P value after Bonferroni adjustment for the three post-baseline time points; P-values <0.05 are shown in bold. Complete pairs, participants with both Week 0 and Week 6 results for a given marker; ITT, all randomized participants with laboratory data. No marker differed between dose groups in any population (all unadjusted P ≥ 0.19). †Log-transformed; Δ is percentage change. ‡UIBC = TIBC − serum iron (derived). Abbreviations: Fe panel, iron and TIBC panel; Tf panel, transferrin panel; other abbreviations as in Table 3.

### 2.4 Modified hair pull test

Hair shedding was evaluated using a researcher-adapted modified hair pull assessment based on the standard clinical hair pull test (24). Participants received standardized written and illustrated instructions and were instructed to grasp a small bundle of hair, approximately the diameter of a marker, from the side of the scalp and apply gentle, steady traction. Extracted hairs were counted and recorded by the participant; lower counts indicate less shedding. The hair shedding endpoint was the change in hair count from Week 0 to Week 6.

### 2.5 Participant-reported outcomes

Participant-reported outcomes were assessed using a researcher-developed 10-item questionnaire of 5-point Likert-scale items grouped into three domains: wellness (Q1, energy; Q2, fatigue; Q3, mood), hair (Q4, shedding; Q5, vibrancy; Q6, perceived new growth; Q7, overall hair quality), and skin (Q8, hydration; Q9, clarity; Q10, overall skin satisfaction). Participants rated their current status for each item. The fatigue and shedding items were reverse-scored so that higher scores indicated more favorable outcomes for every item, and each domain score was calculated as the mean of its items. The full questionnaire is provided in Supplementary Material S1.

### 2.6 Statistical analysis

#### Analysis population

Four analysis populations were defined: intention-to-treat (ITT; all 31 randomized participants analyzed as assigned, 29 with laboratory data), modified ITT (mITT), complete-pairs, and per-protocol. Primary analyses were conducted in the mITT population, instead of the ITT population prespecified in the statistical analysis plan, because of unexpected participant-level deviations, including withdrawal, non-use of the study product, and incomplete blood data (Section 3.1; Figure 1B). One participant who consumed heqLF without iron was retained. Complete-pairs, and per-protocol analyses served as sensitivity analyses.

Complete-pairs analyses of blood biomarkers were conducted within the mITT population and included participants with Week 0 and Week 6 measurements for each marker, without imputation. The blood biomarker per-protocol population required ≥80% compliance, blood draws at Weeks 0 and 6, and no more than one of four draws missed; draws missed because of laboratory error were excused. The modified hair pull test and subjective questionnaire per-protocol populations required ≥80% compliance and completion of both assessments at Weeks 0 and 6. Analyses of these outcomes were restricted to participants with available paired Week 0 and Week 6 data. Missing data were not imputed.

#### Baseline characteristics

Participants’ baseline characteristics were compared between groups using Welch’s unpaired two-sided *t*-tests for continuous variables and Fisher’s exact tests for categorical variables. P-values were not adjusted.

#### Blood biomarkers

Blood biomarkers were analyzed using linear mixed model (LMM) with group, visit (Weeks 0, 2, 4, and 6), and the group × visit interaction as fixed effects, fit by restricted maximum likelihood (REML) with Kenward–Roger degrees of freedom; the residual covariance structure was selected by Akaike’s information criterion and after evaluation of residual Q-Q plots and plots of residual-versus-predicted values.

Ferritin, hs-CRP, and white blood cell count were log-transformed before analysis and are reported as geometric means, with changes expressed as percentages. Ferritin was also adjusted for inflammation using the Biomarkers Reflecting Inflammation and Nutritional Determinants of Anemia (BRINDA) regression approach (57):

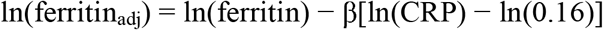

The reference hs-CRP concentration was 0.16 mg/L, based on the BRINDA internal dataset for women of reproductive age (57). The coefficient (β) was estimated separately for each analysis population using a LMM regressing ln(ferritin) on ln(hs-CRP) across visits, with a participant-level random intercept to account for repeated measures (β = 0.153 in the mITT population). Ferritin was adjusted only when hs-CRP was ≥0.16 mg/L; no adjustment was applied when hs-CRP was below this threshold or missing. Because AGP was not measured, adjustment was limited to CRP rather than both CRP and AGP as recommended by BRINDA. Adjusted ferritin values were log-transformed for analysis. hs-CRP values below the limit of detection (LOD) were replaced with LOD/√2.

Within-group changes from Week 0 to Week 6 and between-group differences in these changes were estimated from model contrasts, and pooled changes across both dose groups were estimated from models without a group term. Ferritin was the primary endpoint. P-values were Bonferroni-adjusted for the three post-baseline time points (×3); as a stricter sensitivity analysis, P-values were also adjusted across the 13 blood markers analyzed (×13). Adjusted P-values were capped at 1.

#### Modified Hair pull test

For the modified hair pull test and questionnaire outcomes, normality was assessed using the Shapiro–Wilk test for each dose group (100 mg and 50 mg) at Week 0 and Week 6, and for Week 0–Week 6 change scores, yielding six tests per outcome. As a conservative approach, if at least one Shapiro–Wilk test was significant (*P* < .05), nonparametric methods (Friedman and Wilcoxon signed-rank tests) would be used; otherwise, parametric methods (repeated-measures ANOVA and paired *t*-tests) were used. The alternative approach was used for sensitivity analyses.

Hair pull counts were not normally distributed. Within-group changes from Week 0 to Week 6 were analyzed using Wilcoxon signed-rank tests with continuity correction, with P-values Bonferroni-adjusted for the two dose groups (×2). Changes across weeks were assessed using the Friedman test. Between-group comparisons used the Mann–Whitney *U* test. ANCOVA of Week 6 counts, adjusting for Week 0 counts and including group as a factor, served as the sensitivity analysis. Within-group effect sizes are reported as Cohen’s *d*z (mean change / SD of change , with benchmarks of small (0.2), medium (0.5), and large (0.8) effects.

#### Subjective Questionnaire outcomes

For subjective questionnaire outcomes, within-group changes across Weeks 0–6 within each group was assessed using the Friedman test (Kendall’s *W* as the effect size) in participants with complete data at all seven time points. Each post-baseline week was compared with Week 0 using the Wilcoxon signed-rank test with Bonferroni adjustment for six comparisons (×6). Between groups comparisons at each week used the Mann–Whitney *U* test, with Bonferroni adjustment for seven comparisons (×7).

LMM with group, week, and group × week as fixed effects and a random intercept for participant (REML; Type III *F*-tests with Satterthwaite degrees of freedom) were fit using all available observations. In the mITT population, the non-parametric approach was primary for the wellness and hair domains, and the LMM was primary for the skin domain. Both approaches are reported for all domains.

#### Statistical significance threshold

Statistical significance was set at α = 0.05 (two-sided). Due to the relatively small sample size, the study had no control arm, all analyses are considered exploratory.

## 3 Results

### 3.1 Participant characteristics

Thirty-one women were randomized (100 mg, n = 16; 50 mg, n = 15; Figure 1B). Four were excluded from the mITT population: in the 100 mg group, one did not take the study product, and one had incomplete blood data; in the 50 mg group, one withdrew and one did not take the study product. This left 27 participants (100 mg, n = 14; 50 mg, n = 13). One participant in the 50 mg group who received heqLF without iron was retained. Complete-pairs blood biomarker analyses included 16–23 participants per marker. Per-protocol populations included 18 participants for blood biomarker analyses (100 mg, n = 8; 50 mg, n = 10) and 17 for the hair pull test and questionnaire (100 mg, n = 8; 50 mg, n = 9). Per-protocol results for blood biomarkers and the modified hair pull test are presented in Tables 4 and 5, respectively, and per-protocol results for questionnaire data are presented in Supplementary Table S2.

**Table 5.** Modified hair pull test counts at Week 0 and Week 6.

| Group | n | Week 0, mean (median) | Week 6, mean (median) | Mean change (%) | *d*<sub>z</sub> | Decreased/ increased/ unchanged | P | Adj. P (×2) |
|---|---|---|---|---|---|---|---|---|
| *Modified intention-to-treat* |  |  |  |  |  |  |  |  |
| 100 mg | 11 | 3.64 (3) | 1.36 (1) | −2.27 (−62%) | −0.73 | 10/1/0 | **0.009** | **0.017** |
| 50 mg | 11 | 4.45 (6) | 2.73 (2) | −1.73 (−39%) | −0.96 | 8/1/2 | **0.019** | **0.039** |
| *Per-protocol* |  |  |  |  |  |  |  |  |
| 100 mg | 8 | 4.00 (3) | 1.25 (1) | −2.75 (−69%) | −0.77 | 7/1/0 | **0.029** | 0.058 |
| 50 mg | 9 | 5.00 (6) | 3.22 (3) | −1.78 (−36%) | −0.90 | 6/1/2 | **0.049** | 0.097 |

*Note.* Number of hairs extracted; lower values indicate less shedding. Participants with both Week 0 and Week 6 counts; one participant was excluded for implausible counts. P, Wilcoxon signed-rank test with continuity correction; Adj. P, Bonferroni-adjusted for two dose groups. P-values <0.05 are shown in bold. *d*<sub>z</sub>, mean change ÷ SD of change. Friedman test: 100 mg, *P* = 0.007; 50 mg, *P* = 0.020. Between-group comparison of change (Mann–Whitney *U*): mITT P = 0.95; per-protocol P = 0.85. ANCOVA (Week 6 adjusted for Week 0), 50 mg − 100 mg: mITT +1.08 hairs (95% CI −0.52, +2.68), P = 0.17; per-protocol +1.69 hairs (−0.31, +3.69), P = 0.092.

One internally inconsistent Week 6 iron panel, drawn at an outside laboratory after a missed study draw, was excluded. Two Week 6 ferritin values (429 and 730 ng/mL) were identified as potential outliers based on their magnitude relative to preceding measurements and the absence of evidence of inflammation. The value of 429 ng/mL was excluded from the analysis, while the value of 730 ng/mL was adjusted to 73 ng/mL based on a suspected decimal-place transcription error. One participant was also excluded from hair pull analyses because of implausible counts.

Baseline characteristics of the mITT population are presented in Table 1. Participants were 36.9 ± 9.0 years of age, had a BMI of 25.6 ± 4.6 kg/m², and reported 2.2 ± 2.2 h/wk of resistance training and 4.6 ± 3.3 h/wk of cardiovascular exercise (typical weekly hours, self-reported at baseline). Most participants (73.1%) reported low iron status for more than 1 year prior to enrolling. Mean screening ferritin was 14.0 ± 9.0 ng/mL; 92.6% of participants had screening ferritin <30 ng/mL and 63.0% had values <15 ng/mL. Mean ferritin at Week 0 was higher (31.8 ± 17.6 ng/mL), and 46.2% of participants remained below 30 ng/mL. Three of 24 participants with hemoglobin data (12.5%) were anemic (hemoglobin <12.0 g/dL), and all three met the WHO-based definition of iron-deficiency anemia (hemoglobin <12.0 g/dL with ferritin <15 ng/mL) (56). No baseline characteristic differed significantly between groups (all P ≥ 0.14). Symptom data were available for 25 participants: all reported fatigue, and most reported brain fog (88%) and hair shedding or thinning (84%) at baseline (Table 2).

### 3.2 Iron status, hematological, and inflammatory biomarkers

Changes in blood biomarkers in the mITT population are presented in Table 3. Ferritin did not change significantly from week 0 – week 6 in either group (100 mg: +0.9% [95% CI −18.4, +24.8], P = 0.93; 50 mg: +11.3% [−11.1, +39.4], P = 0.34) or with both groups combined (+5.7% [−9.1, +22.9], P = 0.46), and results were similar after BRINDA adjustment for hs-CRP (AGP not measured).

TIBC and transferrin decreased from week 0 – week 6 in both groups, but only TIBC in the 100 mg group remained significant after adjustment. In the 100 mg group, TIBC significantly decreased by 19 µg/dL (95% CI [−32, −6]; P = 0.005; adjusted P = 0.014) and transferrin by 15 mg/dL (95% CI [−28, −2]; P = 0.020; adjusted P = 0.061). In the 50 mg group, TIBC decreased by 15 µg/dL (95% CI [−29, 0]; P = 0.045; adjusted P = 0.14). Serum iron (100 mg: +13 µg/dL; 50 mg: +15 µg/dL) and TSAT (+4.6 and +5.2 percentage points, respectively) increased in both groups, but these changes were not statistically significant. No between-group differences were detected (all P ≥ 0.41).

From Week 0 to Week 6, pooled TIBC decreased by 17 µg/dL (95% CI [−27, −8]; *P* < 0.001), UIBC by 34 µg/dL (95% CI [−55, −14]; *P* = 0.001), and transferrin by 13 mg/dL (95% CI [−22, −3]; *P* = 0.009). All three decreases remained statistically significant after adjustment (adjusted *P* = 0.002, 0.004, and 0.028, respectively). Pooled serum iron increased by 14 µg/dL (95% CI [−4, +32]) and TSAT increased by 5.0 percentage points (95% CI [−0.2, +10.1]); neither change was statistically significant (all *P* > 0.05). Hemoglobin, hematocrit, hs-CRP, and white blood cell count also showed no significant changes in either group (all *P* > 0.05).

Findings were broadly consistent across analysis populations (Table 4). In the complete-pairs population, pooled TSAT (Fe panel) increased by 6.7% (95% CI [+1.5%, +11.9%]; *P* = 0.015; adjusted *P* = 0.044), and UIBC decreased by 37 µg/dL (95% CI [−55, −18]; adjusted *P* = 0.002); both remained significant after adjustment. Serum iron increased by 22 µg/dL (95% CI [+4, +40]; *P* = 0.017), but this change was not significant after adjustment (adjusted *P* = 0.052). In the per-protocol population, TSAT (Fe panel) increased by 7.3% (95% CI [+1.5%, +13.0%]; adjusted *P* = 0.042), while TIBC decreased by 13 µg/dL (95% CI [−23, −4]; adjusted *P* = 0.025) and UIBC decreased by 38 µg/dL (95% CI [−60, −16]; adjusted *P* = 0.004). Serum iron increased by 22 µg/dL (95% CI [+2, +42]; *P* = 0.028), but this change was not significant after adjustment (adjusted *P* = 0.085). In the ITT population, only the pooled decrease in UIBC remained significant after adjustment (−33 µg/dL; 95% CI [−48, −18]; adjusted *P* < 0.001); the decrease in TIBC did not reach statistical significance after adjustment (adjusted *P* = 0.051). Ferritin did not change significantly, and no marker differed between doses in any population. Replacing the implausible Week 6 ferritin value with 73 ng/mL rather than excluding it did not change these conclusions.

### 3.3 Hair shedding

Results of the modified hair pull test are presented in Table 5. Counts at both Week 0 and Week 6 were available for 11 participants in each group. In the 100 mg group, the mean count significantly decreased from 3.64 (median 3) to 1.36 (median 1) hairs, a reduction of 2.27 hairs (−62%; *d*_z_ = −0.73; P = 0.009; adjusted P = 0.017), indicating a large effect. In the 50 mg group, the mean count significantly decreased from 4.45 (median 6) to 2.73 (median 2) hairs, a reduction of 1.73 hairs (−39%; *d*_z_ = −0.96; P = 0.019; adjusted P = 0.039), indicating a large effect.

Friedman tests indicated significant change across weeks in both groups (100 mg, P = 0.007; 50 mg, P = 0.020). The reduction did not differ between groups (Mann–Whitney P = 0.95), and the ANCOVA sensitivity analysis showed no significant group difference in Week 6 counts after adjustment for baseline (50 mg − 100 mg: +1.08 hairs [95% CI −0.52, +2.68]; P = 0.17). In the per-protocol population, reductions were of similar magnitude (100 mg: −2.75 hairs, −69%; 50 mg: −1.78 hairs, −36%) but did not reach significance after adjustment (adjusted P = 0.058 and 0.097, respectively).

### 3.4 Participant-reported outcomes

Questionnaire results for the mITT population are presented in Table 6. Scores in all three domains changed significantly across Weeks 0–6 within both groups (Friedman P ≤ 0.011; Kendall’s *W* = 0.39–0.85). In the 100 mg group, wellness scores increased from 2.36 at Week 0 to 3.90 at Week 6 (adjusted P = 0.015), hair scores from 1.96 to 3.77 (adjusted P = 0.015), and skin scores from 2.28 to 3.72 (adjusted P = 0.023). In the 50 mg group, wellness scores increased from 2.44 to 3.72 (adjusted P = 0.023; significant at Weeks 3, 4, and 6) and hair scores from 2.21 to 3.33 (adjusted P = 0.035; significant at Week 6 only), whereas the increase in skin scores (2.92 to 3.72) was not significant (adjusted P = 0.145).

**Table 6.**
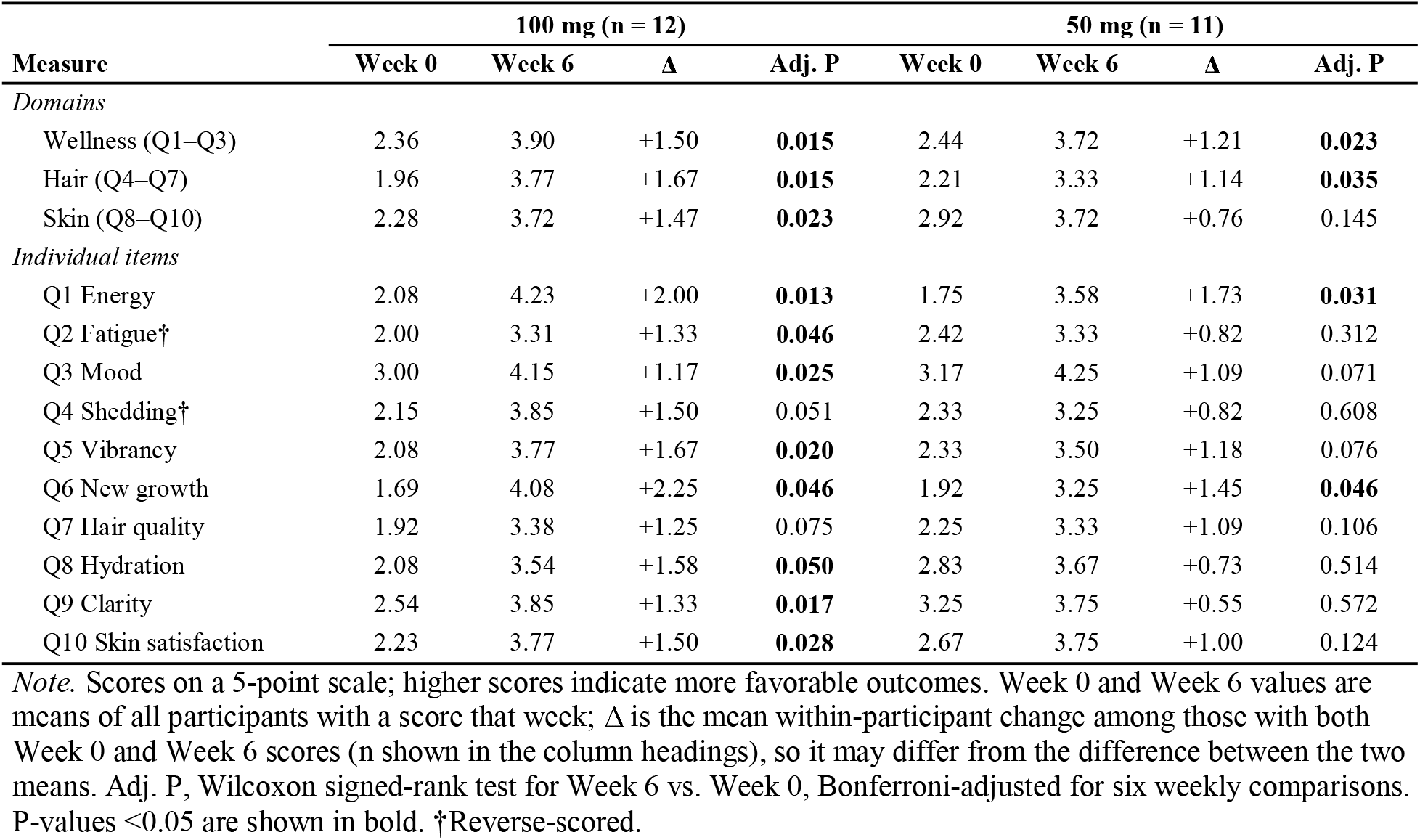
Participant-reported outcomes at Week 0 and Week 6 (modified intention-to-treat).

Domain scores did not differ between groups at any week (Mann–Whitney adjusted P ≥ 0.34). In the mixed-model analyses, the effect of time was significant for all domains (P < 0.001), and the group × time interaction was significant for the skin domain (the primary analysis for this domain; P = 0.009) and the hair domain (P = 0.039), but not for wellness (P = 0.17), indicating that the trajectories of perceived hair and skin health over time differed between doses.

At the item level, energy improved significantly by Week 6 in both groups (100 mg: 2.08 to 4.23, adjusted P = 0.013; 50 mg: 1.75 to 3.58, adjusted P = 0.031), as did perceived new hair growth (100 mg: 1.69 to 4.08; 50 mg: 1.92 to 3.25; both adjusted P = 0.046). In the 100 mg group, fatigue (adjusted P = 0.046), mood (P = 0.025), hair vibrancy (P = 0.020), skin clarity (P = 0.017), overall skin satisfaction (P = 0.028), and skin hydration (P = 0.050) improved significantly. Changes in self-perceived shedding and overall hair quality were not statistically significant (both *P* > 0.05). In the 50 mg group, the remaining items were not significant by weeks (all *P* > 0.05). No item differed significantly between groups at any week (all adjusted *P* > 0.05).

In the per-protocol population, the direction and magnitude of change were similar, but Week 6 improvements in domain scores were not significant after adjustment (adjusted P = 0.054–0.309). Friedman tests remained significant for all domains within both groups (P ≤ 0.007) except skin in the 50 mg group (P = 0.051), and the group × time interaction was significant for the hair (P < 0.001) and skin (P = 0.049) domains, for which the mixed model was the primary analysis in this population. Full per-protocol questionnaire results are provided in Supplementary Table S2.

## 4 Discussion

To our knowledge, this is the first randomized, double-blind study of heqLF administered with iron in women with low ferritin. Over 6 weeks, both doses (50 and 100 mg/d, each with 5 mg/d iron) were accompanied by decreases in TIBC and transferrin, with smaller increases in serum iron and TSAT, whereas ferritin, the primary endpoint, did not change. Hair pull counts decreased by 39–62%, participant-reported wellness and perceived hair health improved in both groups, and perceived skin health improved significantly only with 100 mg. Responses were similar with both doses. The consistency of the iron transport findings across analysis populations, together with parallel improvements in hair shedding and participant-reported outcomes, provides an encouraging signal that warrants confirmation in a controlled trial. Below, we interpret these findings in the context of previous studies of iron and LF supplementation.

### 4.1 Iron transport markers and ferritin

Hepatic transferrin synthesis is upregulated when iron stores are depleted, so transferrin and TIBC are elevated in iron deficiency and fall as iron supply to tissues improves (1, 2). The pooled decreases observed here in TIBC (−17 µg/dL), transferrin (−13 mg/dL), and UIBC (−34 µg/dL) are therefore consistent with improved iron availability. These changes were directionally consistent across all four analysis populations, and the decrease in UIBC remained significant after adjustment in each. Increases in TSAT (+6.7 to +7.3 percentage points) and serum iron (+22 µg/dL) were nominally significant in the complete-pairs and per-protocol analyses but not in the mITT or ITT analyses, suggesting that missing visits attenuated these signals. Because transferrin has a circulating half-life of roughly 8 days, transport markers can respond within weeks, whereas a measurable rise in ferritin requires sustained net iron retention. A response in transport markers without a change in ferritin is therefore plausible over a 6-week period. UIBC is derived from TIBC and serum iron and does not provide independent evidence.

The stable ferritin concentrations are best understood by comparison with iron trials in similar populations. In non-anemic menstruating women with fatigue and ferritin <50 µg/L, 80 mg/d of elemental iron for 12 weeks raised ferritin by 11.4 µg/L and hemoglobin by 0.32 g/dL relative to placebo (8). The present study provided 16-fold less iron for half as long. Stable-isotope studies in iron-depleted women show that fractional absorption is highest at low doses and declines once doses reach ≥60 mg, because such doses increase hepcidin for about 24 h; absolute absorption, however, still rises with dose, from 6.7 mg at a 40 mg dose to 18.1 mg at 240 mg (28, 29). Even with efficient absorption, a 5 mg dose is therefore likely to supply no more than 1–2 mg of absorbed iron per day, which is similar in magnitude to the basal requirement of menstruating women (2) and leaves little surplus for rebuilding stores within 6 weeks. The well-absorbed iron form may have partly offset this: ferrous bisglycinate improves hemoglobin more than other iron salts in pregnant women and caused fewer gastrointestinal adverse events (58). In addition, mean ferritin at Week 0 (31.8 ng/mL) was more than twice the mean screening value (14.0 ng/mL), and nearly half of participants had Week 0 ferritin ≥30 ng/mL, which narrowed the scope for improvement. This gap may reflect regression to the mean, iron intake between screening and enrollment, or inter-laboratory differences, because screening values came from participants’ own records. Inflammation is unlikely to have masked a ferritin response: hs-CRP was low and stable, and BRINDA-adjusted ferritin followed the same pattern (57).

### 4.2 Comparison with previous lactoferrin studies

Most clinical evidence for LF comes from bLF in pregnant women with ID or IDA. In those studies, bLF increased hemoglobin, serum iron, and ferritin and decreased IL-6, whereas ferrous sulfate increased IL-6 (39, 42). Meta-analyses have concluded that bLF is at least as effective as ferrous sulfate for hematological recovery in pregnancy, with fewer gastrointestinal side effects (43), and that it produces larger gains in ferritin despite lower fractional iron absorption (44). Several features distinguish those studies from the present one. Participants in previous bLF studies were generally anemic or more severely iron-deficient, LF was typically given at 200 mg/d (100 mg twice daily) or more, and responses were often driven by correction of inflammation-related iron sequestration (38, 39). In contrast, our participants were largely non-anemic but experiencing absolute iron deficiency with low baseline hs-CRP (mean 2.1 mg/L) that did not change, so other mechanisms, such as receptor-mediated uptake of iron-bound LF (36, 37), may be more relevant in this population.

A recent large trial helps clarify the role of LF. In 555 Bangladeshi women with IDA, bLF at 200 or 400 mg/d for 12 weeks, given without iron, was inferior to 60 mg/d ferrous sulfate for hemoglobin and increased ferritin by only about 3 µg/L, compared with about 41 µg/L with ferrous sulfate; CRP and α1-acid glycoprotein did not differ between groups (40). Notably, hepcidin rose only minimally with bLF (0.5–1.0 ng/mL) compared with about 16 ng/mL with ferrous sulfate (40). These findings suggest that LF is best positioned as a complement to iron rather than a replacement, and the absence of a hepcidin surge makes it an attractive adjunct to optimize fractional iron absorption with low-dose iron, the approach tested here. Because ferrous sulfate also raised ferritin far more, its larger hepcidin response may partly reflect greater iron repletion; the proposed benefit of LF therefore remains enhanced fractional iron absorption, which was not measured here. This combination is also supported by a study of 16 female long-distance runners, in which ferritin, serum iron, and red blood cell count declined during 8 weeks of summer training with iron alone but were maintained with LF plus iron (45). The present study is consistent with these findings and extends them to free-living women with low ferritin and to a heqLF, which is digested more like human milk LF than bLF is (52) and is the native ligand of the intestinal LF receptor (36, 47). However, without an iron-only control arm and with no change in ferritin, the present study cannot isolate an effect of LF.

In the case series that motivated the present study, 100 mg/d heqLF was followed by marked increases in ferritin in two active women with persistently low iron over 6–20 weeks (54), suggesting that longer exposure may allow effects on iron transport to translate into replenished stores. Contrary to our hypothesis, changes were similar with 50 and 100 mg/d, indicating that the lower dose may be sufficient for effects on iron transport; larger samples will be needed to define the dose–response relationship, as between-group confidence intervals were wide (for ferritin, −19% to +50%). Both doses were also far below the 0.34 and 3.4 g/d evaluated in a 28-day safety trial in healthy adults (53).

### 4.3 Hair shedding

Hair pull counts decreased by 2.3 hairs (−62%) with 100 mg and 1.7 hairs (−39%) with 50 mg, with moderate-to-large within-group effect sizes (*d*_z_ −0.73 and −0.96), and 10 of 11 and 8 of 11 participants, respectively, shed fewer hairs at Week 6. Evidence linking iron status to hair shedding is mixed: case-control studies have reported lower ferritin in women with diffuse non-scarring hair loss (15–18), but larger controlled studies found no excess of ID or no association above a ferritin of 10 µg/L (19, 20), and interventional evidence is sparse (6, 21). Because ferritin did not change, the reduction in shedding may involve mechanisms beyond iron stores. An iron-independent action of LF is biologically plausible: LF promoted hair growth in mice and stimulated dermal papilla cell proliferation through ERK/Akt and Wnt signaling (22), although whether orally ingested LF or its peptides reach the follicle remains to be determined. A recent review identified no registered human trials of LF for hair outcomes (23), so the present data are, to our knowledge, the first clinical observations of hair shedding during LF supplementation.

Reductions of similar magnitude were observed in the per-protocol population. As in any uncontrolled comparison, seasonal variation, regression to the mean, and growing familiarity with the self-administered test may also have contributed, so confirming these encouraging findings with clinician-performed pull tests (24) or phototrichograms in a placebo-controlled trial will be an important next step.

### 4.4 Participant-reported outcomes

Energy improved in both groups by 1.7–2.0 points on a 5-point scale, and the 100 mg group also reported improvements in fatigue and mood. These changes are directionally consistent with placebo-controlled iron trials in non-anemic women. In those trials, fatigue fell by 47.7% with iron vs. 28.8% with placebo over 12 weeks (8) and by 29% vs. 13% over 4 weeks, with benefit limited to women with ferritin ≤50 µg/L (9). A systematic review likewise found that iron reduced subjective fatigue in non-anemic iron-deficient adults without improving objective physical capacity (10). Because placebo groups in these trials also improved, and our questionnaire was researcher-developed, expectation effects may have contributed, and validated instruments with a placebo comparator will help quantify the specific contribution of heqLF.

The improvements in perceived skin health with 100 mg are consistent with clinical evidence that oral LF reduces inflammatory acne lesions, which may reflect its antimicrobial and anti-inflammatory properties (23). In the mixed models, the significant group × time interactions for the hair and skin domains suggest a steeper trajectory with 100 mg, a pattern worth examining prospectively, although groups did not differ at any individual week.

### 4.5 Analysis populations and multiplicity

Results were examined in several analysis populations, and the rationale warrants explanation. The ITT population as designated as primary, but it included two participants who confirmed they never took the study product and one who withdrew, which would bias estimates toward the null. We therefore present the mITT population, which excludes only participants who did not receive the intervention or contributed no usable blood data, and report the ITT, complete-pairs, and per-protocol analyses as sensitivity analyses so that the robustness of each finding can be judged. The decrease in UIBC remained significant after adjustment in every population. The decrease in TIBC was nominally significant in every population and remained significant after adjustment in two. The increases in serum iron and TSAT were nominally significant only in the complete-pairs and per-protocol analyses, indicating that they depend on how missing data are handled. For the hair pull test and questionnaire, the within-group tests require paired observations, so the mITT analyses of these outcomes are equivalent to complete-pairs analyses; ITT analyses were not performed for these outcomes, which is a limitation.

We applied conservative Bonferroni corrections throughout. Within dose groups, the decreases in UIBC in both groups, in transferrin with 100 mg, and in TIBC with 50 mg were nominally significant but not after Bonferroni adjustment, and in the per-protocol population the hair pull and questionnaire changes were not significant after adjustment. This is expected in a pilot study where 11–14 per group contributed to the main analyses, so these results are best interpreted as estimates of effect size to inform the design of a larger trial.

### 4.6 Strengths and limitations

Strengths of this study include randomized, double-blind allocation to two doses; a decentralized design with mobile phlebotomy that enabled recruitment of community-dwelling women; repeated blood sampling and weekly participant-reported assessments; inflammation-adjusted ferritin; prespecified normality-based analysis rules; and multiple sensitivity analyses.

As a pilot study, this trial also has limitations. Without a placebo or iron-only arm, the specific contribution of heqLF cannot yet be isolated. The sample was small and the intervention short. Some blood data were missing because of laboratory errors and missed draws, and screening ferritin values came from participants’ own records. Hair shedding and participant-reported outcomes were assessed with self-administered instruments that have not yet been validated, and hepcidin and soluble transferrin receptor were not measured.

Missing data were not imputed for the hair pull analysis, reducing the sample size for per-protocol analyses. We chose not to impute missing values for several reasons: (1) we did not have a theoretical or prespecified rationale for selecting predictors for the imputation model; (2) including all available variables without a clear rationale increase the risk of overfitting; (3) the small sample size limited our confidence that the imputed values would reliably represent the actual outcome measures; and (4) given the exploratory, hypothesis-generating nature of the per-protocol analyses, we considered the potential uncertainty introduced by imputation less justifiable than analyzing the available paired data.

### 4.7 Future directions

An adequately powered trial with placebo and iron-only comparator arms, lasting at least 12 weeks, is needed to isolate the effect of heqLF. Such a trial should include hepcidin and soluble transferrin receptor to test the proposed mechanisms, validated fatigue and quality-of-life instruments, and objective measures of hair shedding. Enrolling women with confirmed low ferritin at baseline, rather than at screening, would also increase the scope for detecting changes in iron stores.

## 5 Conclusions

In women with low ferritin, 6 weeks of heqLF at 50 or 100 mg/d with 5 mg/d iron was accompanied by reductions in iron-binding capacity and transferrin (i.e., improved iron availability), fewer hairs shed on a self-administered pull test, and improvements in participant-reported wellness and perceived hair health, without a change in ferritin or differences between doses. These encouraging findings, obtained with lower LF doses and a shorter duration than in previous LF trials, support a larger placebo-controlled trial of heqLF as an adjunct to low-dose iron.

## Data availability statement

The raw data supporting the conclusions of this article will be made available by the authors upon reasonable request.

## Ethics statement

The study involving humans were approved by the BRANY Institutional Review Board (BRANY #26-02-188-2514; Protocol Version 2; approved May 8, 2026).

## Author contributions

Conceptualization: RP, CE. Data curation: CE, MC. Formal analysis: IV, MC, DB. Funding acquisition: RP. Investigation: CE, IV. Methodology: CE, RP. Project administration: CE. Resources: RP. Validation: MC. Visualization: RP. Writing – original draft: IV. Writing – review & editing: IV, CE, MC, DB, RP.

## Acknowledgments

The authors thank the study participants. BioRender (BioRender.com) was used to create figures.

## Generative AI statement

The author(s) verify and take full responsibility for the use of generative AI in the preparation of this manuscript. AI-assisted tools were used for spelling and grammar review. Claude (Anthropic) was used to assist with troubleshooting code used in statistical analysis. AI tools were not used to generate scientific conclusions, interpret study findings, choose statistical analysis, or replace author judgment.

## Supplementary material

The Supplementary Material for this article can be found online at: [link to be added by the publisher]. Supplementary Material S1: participant-reported outcome questionnaire. Supplementary Table S2: per-protocol results for questionnaire data.

