## Supplementary Material S1 for "Effects of human lactoferrin combined with low-dose iron on iron status, hair and skin in women: a randomized, double-blind pilot study"

**Supplementary Material S1.** Participant-reported outcome questionnaire (Part A) and modified hair pull test instructions (Part B) provided to participants.

**Part A. Wellness, hair and skin survey**

**Instructions**

Please answer the following questions based on the last 7 days.

Scale:

1 = Strongly Disagree

2 = Disagree

3 = Neutral

4 = Agree

5 = Strongly Agree

*Mood and Energy*

1.	I have felt energetic throughout the day.

2.	I have felt physically fatigued during the day.

3.	My overall mood has been positive.

*Hair*

4.	I am experiencing hair shedding

5.	I have noticed an improvement in overall hair quality

6.	My hair appears shiny and vibrant

7.	I have noticed new hairs growing in

*Skin*

8.	My skin feels hydrated

9.	My skin appears clear

10.	I have noticed an improvement in overall skin health

11.	I am experiencing dry skin

**Part B. Hair pull test instructions**

**Hair Pull Test Instructions**

The hair pull test is a simple way to measure how much hair is shedding.

**Before You Begin**

- Do **not wash your hair for at least 24 hours** before completing this test.
- Perform the test on **dry hair**.
- Make sure your **hands are clean and dry**.

**Step 1: Select a Small Section of Hair**

Using your fingers, separate a **small bundle of hair from the top of your scalp (crown area)**.

The bundle should be about the **width of a small pencil** (about 20–60 hairs).

**Step 2: Hold the Hair**

Grip the hair **close to the scalp** using your **thumb, index finger, and middle finger**.

**Step 3: Gently Pull**

Firmly but gently **pull the hair away from the scalp** while sliding your fingers down toward the ends of the hair.

- The pull should be **firm but not painful**.
- You may feel **slight tension on the scalp**, but it should not hurt.

**Step 4: Count the Hairs**

Look at the hairs that came out during the pull and **count how many strands were removed**.

**Step 5: Record Your Result**

Write down the **number of hairs that came out during the test**.
