## Supplementary Table S2 for "Effects of human lactoferrin combined with low-dose iron on iron status, hair and skin in women: a randomized, double-blind pilot study"

**Supplementary Material**

**Supplementary Table S2.** Participant-reported outcomes at Week 0 and Week 6 (per-protocol population).

|  | **100 mg (n = 8)** | | | | **50 mg (n = 9)** | | | |
| --- | --- | --- | --- | --- | --- | --- | --- | --- |
| **Measure** | **Week 0** | **Week 6** | **Δ** | **Adj. P** | **Week 0** | **Week 6** | **Δ** | **Adj. P** |
| *Domains* | | | | | | | | |
| Wellness (Q1–Q3) | 2.46 | 4.00 | +1.54 | 0.085 | 2.48 | 3.70 | +1.22 | 0.054 |
| Hair (Q4–Q7) | 1.78 | 3.81 | +2.03 | 0.084 | 2.36 | 3.28 | +0.92 | 0.085 |
| Skin (Q8–Q10) | 2.25 | 3.75 | +1.50 | 0.134 | 3.00 | 3.67 | +0.67 | 0.309 |
| *Individual items* | | | | | | | | |
| Q1 Energy | 2.12 | 4.12 | +2.00 | 0.080 | 1.78 | 3.56 | +1.78 | 0.077 |
| Q2 Fatigue† | 2.25 | 3.75 | +1.50 | 0.186 | 2.56 | 3.33 | +0.78 | 0.572 |
| Q3 Mood | 3.00 | 4.12 | +1.12 | 0.106 | 3.11 | 4.22 | +1.11 | 0.201 |
| Q4 Shedding† | 2.25 | 3.75 | +1.50 | 0.124 | 2.67 | 3.11 | +0.44 | 1.000 |
| Q5 Vibrancy | 2.00 | 3.75 | +1.75 | 0.126 | 2.44 | 3.44 | +1.00 | 0.192 |
| Q6 New growth | 1.25 | 4.38 | +3.12 | 0.071 | 1.89 | 3.22 | +1.33 | 0.106 |
| Q7 Hair quality | 1.62 | 3.38 | +1.75 | 0.119 | 2.44 | 3.33 | +0.89 | 0.320 |
| Q8 Hydration | 2.00 | 3.38 | +1.38 | 0.341 | 2.89 | 3.56 | +0.67 | 1.000 |
| Q9 Clarity | 2.50 | 3.88 | +1.38 | 0.119 | 3.44 | 3.89 | +0.44 | 1.000 |
| Q10 Skin satisfaction | 2.25 | 4.00 | +1.75 | 0.107 | 2.67 | 3.56 | +0.89 | 0.339 |

Per-protocol population (≥80% compliance; questionnaire completed at Weeks 0 and 6). Scores on a 5-point scale; higher scores indicate more favorable outcomes. Week 0 and Week 6 values are means of all participants with a score that week; Δ is the mean within-participant change among those with both Week 0 and Week 6 scores (n shown in the column headings), so it may differ from the difference between the two means. Adj. P, Wilcoxon signed-rank test for Week 6 vs. Week 0, Bonferroni-adjusted for six weekly comparisons. P-values <0.05 would be shown in bold; no Week 6 adjusted P-value met this threshold in this population. Between-dose comparisons at each week (Mann–Whitney U, Bonferroni-adjusted for seven comparisons) were not significant for any domain (adjusted P ≥ 0.73) or item (adjusted P ≥ 0.37). †Reverse-scored.

**Supplementary Table S2 (continued).** Within-group change across weeks and group × time effects for questionnaire domain scores (per-protocol population).

|  |  | **Friedman, 100 mg (N = 6)** | | | **Friedman, 50 mg (N = 6)** | | | **Group × time** | |
| --- | --- | --- | --- | --- | --- | --- | --- | --- | --- |
| **Domain** | **Primary analysis** | **χ²** | **P** | ***W*** | **χ²** | **P** | ***W*** | **F (df)** | **P** |
| Wellness (Q1–Q3) | Non-parametric | 28.57 | **<0.001** | 0.79 | 21.23 | **0.002** | 0.59 | 1.20 (6, 85.3) | 0.312 |
| Hair (Q4–Q7) | Mixed model | 30.69 | **<0.001** | 0.85 | 17.68 | **0.007** | 0.49 | 6.02 (6, 85.2) | **<0.001** |
| Skin (Q8–Q10) | Mixed model | 22.65 | **<0.001** | 0.63 | 12.53 | 0.051 | 0.35 | 2.21 (6, 85.2) | **0.049** |

Per-protocol population. Friedman test of change across Weeks 0–6 within each dose group, in participants with complete data at all seven time points (N in column headings); W, Kendall’s coefficient of concordance (effect size). Group × time, F-test for the group × week interaction from a linear mixed model with group, week, and group × week as fixed effects and a random intercept for participant (REML; Type III F-tests with Satterthwaite degrees of freedom), fit using all available observations; degrees of freedom are numerator, denominator. The main effect of week was significant for all three domains (P < 0.001), and the main effect of group was not (P ≥ 0.75). Primary analysis, the analysis designated as primary for each domain according to the statistical analysis plan. P-values <0.05 are shown in bold.
